# Multi-faceted analysis of COVID-19 epidemic in the Republic of Korea considering Omicron variant: Mathematical modeling-based study

**DOI:** 10.1101/2022.04.15.22273907

**Authors:** Youngsuk Ko, Victoria May Mendoza, Renier Mendoza, Yubin Seo, Jacob Lee, Jonggul Lee, Donghyok Kwon, Eunok Jung

**Author notes:** **Address for Correspondence:** Eunok Jung, PhD, Department of Mathematics, Konkuk University, Seoul, Korea.

## Abstract

**Background:** The most recent variant of concern, Omicron (B.1.1.529), has caused numerous cases worldwide including the Republic of Korea due to its fast transmission and reduced vaccine effectiveness.

**Methods:** A mathematical model considering age-structure, vaccine, antiviral treatment, and influx of the Omicron variant was developed. We estimated transmission rates among age groups using maximum likelihood estimation for the age-structured model. The impact of nonpharmaceutical interventions (in community and border), quantified by a parameter *μ*in the force of infection, and vaccination were examined through a multi-faceted analysis. A theory-based endemic equilibrium study was performed to find the manageable number of cases according to Omicron-and healthcare-related factors.

**Results:** By fitting the model to the available data, the estimated values of *μ* ranged from 0.31 to 0.73, representing the intensity of nonpharmaceutical interventions such as social distancing level. If *μ* < 0.55 and 300,000 booster shots were administered daily from February 3, 2022, the number of severe cases was forecasted to exceed the severe bed capacity. Moreover, the number of daily cases is reduced as the timing of screening measures is delayed. If screening measure was intensified as early as November 24, 2021 and the number of overseas entrant cases was contained to 1 case per 10 days, simulations showed that the daily incidence by February 3, 2022 could have been reduced by 87%. Furthermore, we found that the incidence number in mid-December 2021 exceeded the theory-driven manageable number of daily cases.

**Conclusion:** Nonpharmaceutical interventions, vaccination, and antiviral therapy influence the spread of Omicron and number of severe cases in the Republic of Korea. Intensive and early screening measures during the emergence of a new variant is key in controlling the epidemic size. Using the endemic equilibrium of the model, a formula for the manageable daily cases depending on the severity rate and average length of hospital stay was derived so that the number of severe cases does not surpass the severe bed capacity.

## INTRODUCTION

The severe acute respiratory syndrome coronavirus 2 (SARS-CoV-2) disease (COVID-19), which originated in China at the end of 2019, has become a global public health issue and was declared a pandemic by the World Health Organization (WHO) on 11 March 2020.^1,2^ In mid-November 2021, a new variant, called Omicron, was detected in Gauteng province, South Africa. ^3^ On November 26, 2021, Omicron variant was designated by the Technical Advisory Group on SARS-CoV-2 Virus Evolution of WHO as a variant of concern (VOC). ^4^ In sample serums from vaccinated individuals, the neutralization of Omicron variant was much less compared to the previous variants. ^5^ Moreover, the vaccine effectiveness of primary dose was shown to be reduced against symptomatic Omicron infections. ^6,7^ Vaccine-breakthrough Omicron infections are higher when compared to Delta. ^8^ On the other hand, reduced hospitalization rates and fewer severe cases are observed. ^9,10^ Vaccination remains a key intervention strategy as it offers protection against hospitalization. ^6^ Furthermore, booster shots can provide a substantial increase in protection against symptomatic. ^6,7^ The development of safe and effective oral antiviral drugs can significantly impact control measures for COVID-19. ^11^ In particular, Pfizer’s Paxlovid has been shown to be 89% effective in reducing the risk of hospitalization. ^12^

In Korea, Omicron variant cases have been detected since November 2021 and later, Omicron variant has become the dominant strain, reaching over 50% in mid-January 2022 and more than 90% among confirmed cases since February 2022. ^13^ After the Omicron variant became dominant, the number of cases increased significantly. Average daily confirmed cases in December 2021 (Delta-dominant) and March 2022 (Omicron-dominant) were approximately 6,000 and 300,000, respectively. Since 10 February 2022 and 21 February 2022, the antiviral pill Paxlovid has been given to infected individuals over 60 year and over 40 years, respectively. ^14,15^

Mathematical modelling has been extensively used throughout the different phases of the pandemic. During the early stage of COVID-19, mathematical models were used to forecast the number of cases in various countries. ^16-19^ Non-pharmaceutical interventions (NPIs) such as massive testing, contact tracing, social distancing, mobility restrictions, school closure, mask mandate, etc., have been incorporated in models to come up with effective policies in curbing the rise of infections. ^21-25^ Strategies for vaccine rollout were also proposed using mathematical models. ^25-27^ Because variants may have different epidemiological characteristics, they have been incorporated into models to capture their dynamics and propose strategies to mitigate their spread. ^28,29^

In our proposed mathematical model, we considered the Delta and Omicron variants. We incorporated age structure, foreign entrant cases, vaccination, and antiviral therapy in the model. The aim of this study is to quantify and analyze the impacts of NPIs, such as social distancing and screening measures at the border, in controlling the spread of the disease. Furthermore, we forecasted the number of daily incidence and severe infections caused by the Omicron variant in the Republic of Korea in 2022. By analyzing the endemic equilibrium of the model, we determined the number of manageable daily cases so that the number of severe cases will not surpass the severe bed capacity in Korea.

## METHODS

### Mathematical modeling of COVID-19 considering Delta and Omicron variants

We consider eight age groups and two strains of COVID-19, Delta (*δ*) and Omicron (*o*). These variants have different basic reproductive numbers, transmission rates, and severe rates. Transmission rates among age groups were estimated using maximum likelihood estimation (MLE). The description of the estimation can be found in the Appendix. Fig.1 illustrates the flow diagram of the mathematical model. Note that *X* indicates vaccine- or waning-related status of the host and *i* refers to age group. There are seven vaccine- or waning-related compartments (*X*); *u* (unvaccinated), *w* (unvaccinated, previously infected, but natural immunity has waned), *v1* (two weeks before finishing primary doses), *v2* (two weeks after finishing primary doses), *wv* (waned after primary doses), *b* (boostered), *wb* (waned after booster).

**Fig.1.**
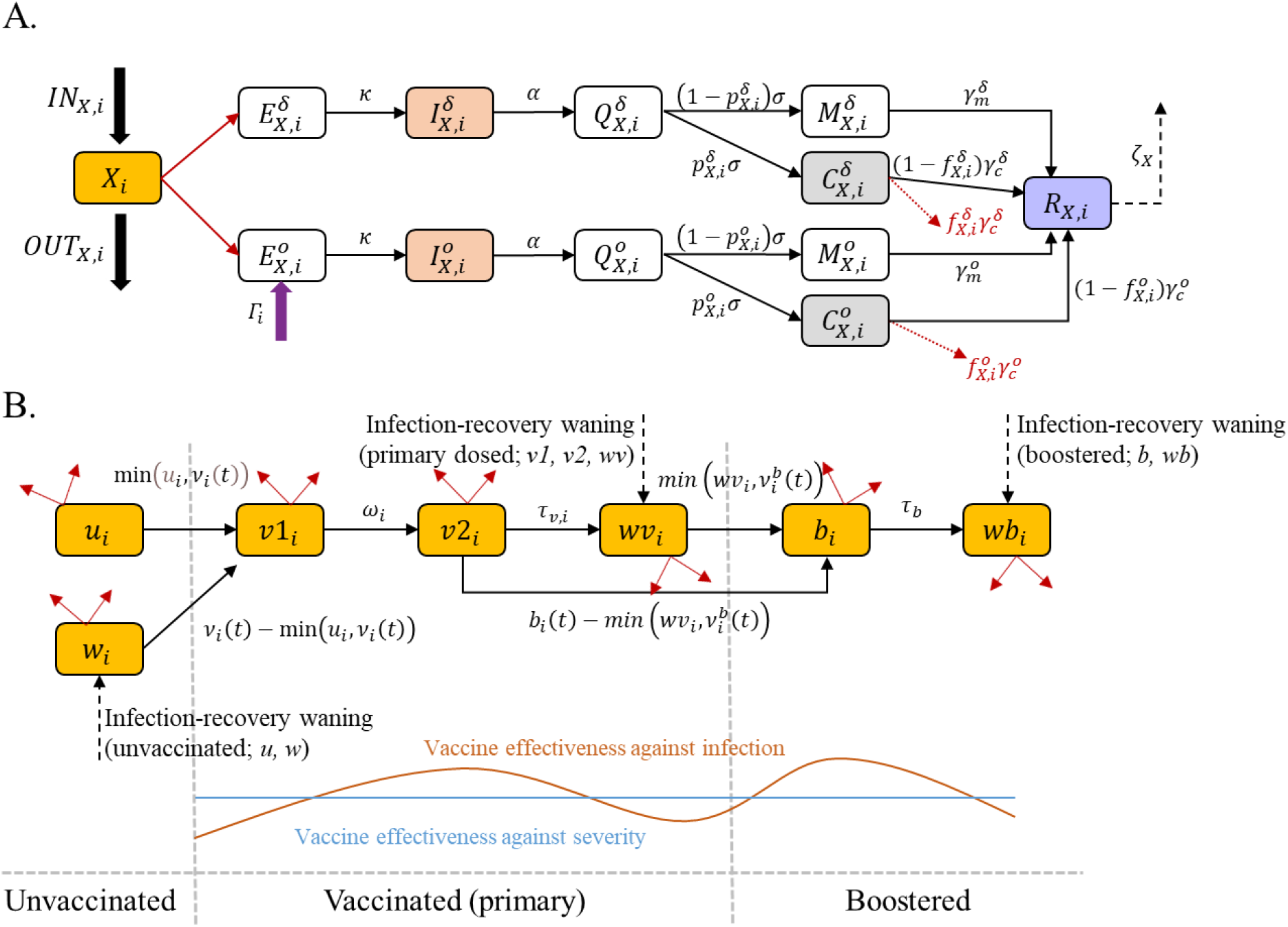
Flow diagrams of the mathematical model of COVID-19 in Korea. (A) Epidemiological flow diagram, where *X*_*i*_ represents a vaccine-or waning-related status of a host in compartment *X* and age group *i*. Note that *X* can be *u, w, v1, v2, wv, b*, or *wb* and each follows this epidemiological flow. (B) Flow diagram describing vaccination, including booster, and waning of immunity after vaccination or infection, which constitute the IN flow to and OUT flow from each *X*_*i*_ in (A). The time-dependent parameters *v*_*i*_(*t*) and 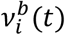 are the number of primary and booster doses administered per day and are obtained from data. The blue line in the bottom graph shows that the values used for vaccine effectiveness against severe infection are the same across all vaccinated individuals but the vaccine effectiveness against infection (red curve) peaks after completing the primary dose and after getting a booster shot.

An unvaccinated host (*u*_*i*_) moves to the *ν*1_*i*_ compartment after administration of the first dose and has partial vaccine effectiveness. Two weeks after receiving the second dose, the host has full vaccine effectiveness (*ν*2_*i*_). Later, vaccine-induced immunity wanes and so the host moves to the *wν*_*i*_ compartment. The host goes to *b*_*i*_ compartment after receiving a booster shot and later to *wb*_*i*_, considering the waning of booster shots. In this study, we assume that the immunity against symptomatic infection wanes but the immunity against severe infection does not. This assumption is supported by studies, where a population-wide decline in effectiveness against infection was observed but effectiveness against hospitalization remained high and with no significant change over time. ^30,31^

For both variants, an exposed host *E*_*X,i*_ becomes infectious (*I*_*X,i*_) and spreads the disease until case confirmation, and so the host moves to the *Q*_*X,i*_ compartment. After confirmation, the isolated host either develops mild symptoms *M*_*X,i*_, including asymptomatic case, or severe/critical symptoms *C*_*X,i*_. An isolated host with mild symptoms recovers (*R*_*X,i*_), while those with severe symptoms may either recover or die. We assume that recovered individuals, whether vaccinated or not, develop natural immunity which wanes over time. Since recovered individuals retain protection against severe infection, they move to a different compartment *w*_*i*_ for unvaccinated, *wν*_*i*_ for primary-dosed, or *wb*_*i*_ for boostered, after the natural immunity has waned. ^31^ It was demonstrated that in unvaccinated participants, the infection-acquired immunity waned after about 1 year but remained consistently high in previously vaccinated-participants, even for individuals who were infected 18 months prior. ^32^ Hence, we use a different natural immunity waning rate for those who were unvaccinated (*ζ*) and vaccinated (*ζ*_*ν*_), with *ζ*_*ν*_< *ζ*. The parameter Γ_*i*_ represents the number of overseas entrant cases from age group *i* who are not screened and entered the local community. Its value is calculated using data on average daily number of overseas entrant cases across all ages from November 24 to December 31, 2021. The detailed description of the mathematical model, including the governing equations, can be found in the Appendix.

To quantify the impact of NPIs, a time-dependent parameter *μ* is introduced to indicate the reduction in transmission caused by NPIs. For example, ignoring other factors, if the basic reproductive number is 2 and *μ* is 0.7, then the effective reproductive number becomes (1 − 0.7) × 2 = 0.6. We estimated the value of *μ* every week using least squares curve fitting method, by minimizing the difference between the cumulative incidence calculated using the model 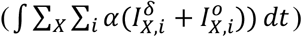 and the available data. The model simulation time was done from August 1, 2021 to February 2, 2022, because the testing policy has been changed since February 3, 2022. ^33^ To apply impact of antiviral therapy, we simply set that severity rate of age over 60 and 40 reduce since January 14 and February 21, 2022, by 80%, respectively. ^14,15^

In this research, we performed a multifaceted approach to examine critical factors which affected the COVID-19 epidemic. First, we did a forecast considering different NPIs-related factors and vaccine hesitancy. Second, we examined the time-dependent sensitivity of screening measures to the disease spread since the Omicron variant has arrived in Korea. Finally, we derived a manageable daily incidence number from the endemic equilibrium state of the mathematical model.

### Forecast of Omicron variant epidemic in 2022

For the forecast, we extended the simulation time until the end of 2022 and varied the factors related to NPIs and booster shots. We set the range for the NPIs-related reduction factor (*μ*) from 0.4 to 0.65 in 0.05 increments (six scenarios), and the maximum number of daily booster shots as 300,000 or 100,000, to observe the influence of vaccine hesitancy. Furthermore, to consider the implementation of the antiviral therapy, we set that groups of age over 60 (40) have reduced severity after 10 February 2022 (21 February 2022).

### Examination of the time-dependent impact of screening measure

We examined the impact of screening measures by varying the value of Γ_*i*_ by factors of 0.1 to 10, and the date of entry of Omicron to the local community from Nov-24 to Dec-1, Dec-8, and Dec-22. The rest of the parameters are fixed to their values on Table^a^ and Table^b^ in the Appendix.

### Endemic equilibrium study

As the number of cases rapidly increases, endemicity of COVID-19 becomes an issue. We performed an endemic equilibrium analysis to investigate how COVID-19 cases can be maintained on a manageable level. Ignoring age structure (*i*), vaccination-related history (*X*), and strains, and considering endemic equilibrium (assuming that there is natural death and birth in susceptible groups, therefore endemic equilibrium can exist), ODEs of confirmed (*Q*) and severe patients (*C*) are

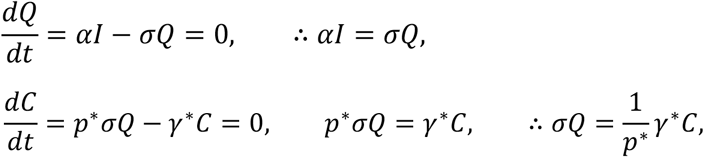

where *p*\* and *γ*\* are average severe rate and recovery rate, respectively. Combining the two results above, we get

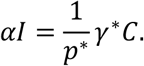

Because the number of severe patients should be below the severe bed capacity, *C*_*max*_, the following inequality is formulated,

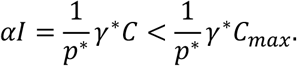

Considering that *αI* is the daily incidence and 1/*γ*\* is the average length of hospital stay, then the threshold value of the inequality, referred to as the manageable daily incidence, is given as follows,

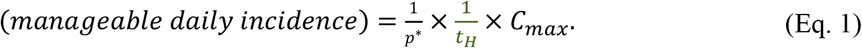

The manageable daily incidence is a function of three input parameters, average length of hospital stay (*t*_*H*_), average severe rate (*p*\*), and severe patient capacity (*C*_*max*_).

### Ethics statement

The study was conducted according to the guidelines of the Declaration of Helsinki, and approved by the Institutional Review Board of Konkuk University (7001355-202101-E-130).

## RESULTS

### Estimation of transmission rates among age groups

Fig.2 shows the transmission rate matrix, *M*_1_, represented as a heatmap. The maximum value is 6.14, which is the value among age group 8. Estimated reproductive number from *M*_1_ is 6.16, which is affected by NPIs but not by vaccine because reduced probability of being infected was considered in the MLE process. To exclude the effect of NPIs, the adjusted matrix *M*_2_ was introduced using the basic reproductive number of the variant and the estimated effective reproductive number from the transmission rate matrix, *M*_1_ (see Appendix). The adjusted matrix *M*_2_ was applied into the mathematical model.

**Fig.2.**
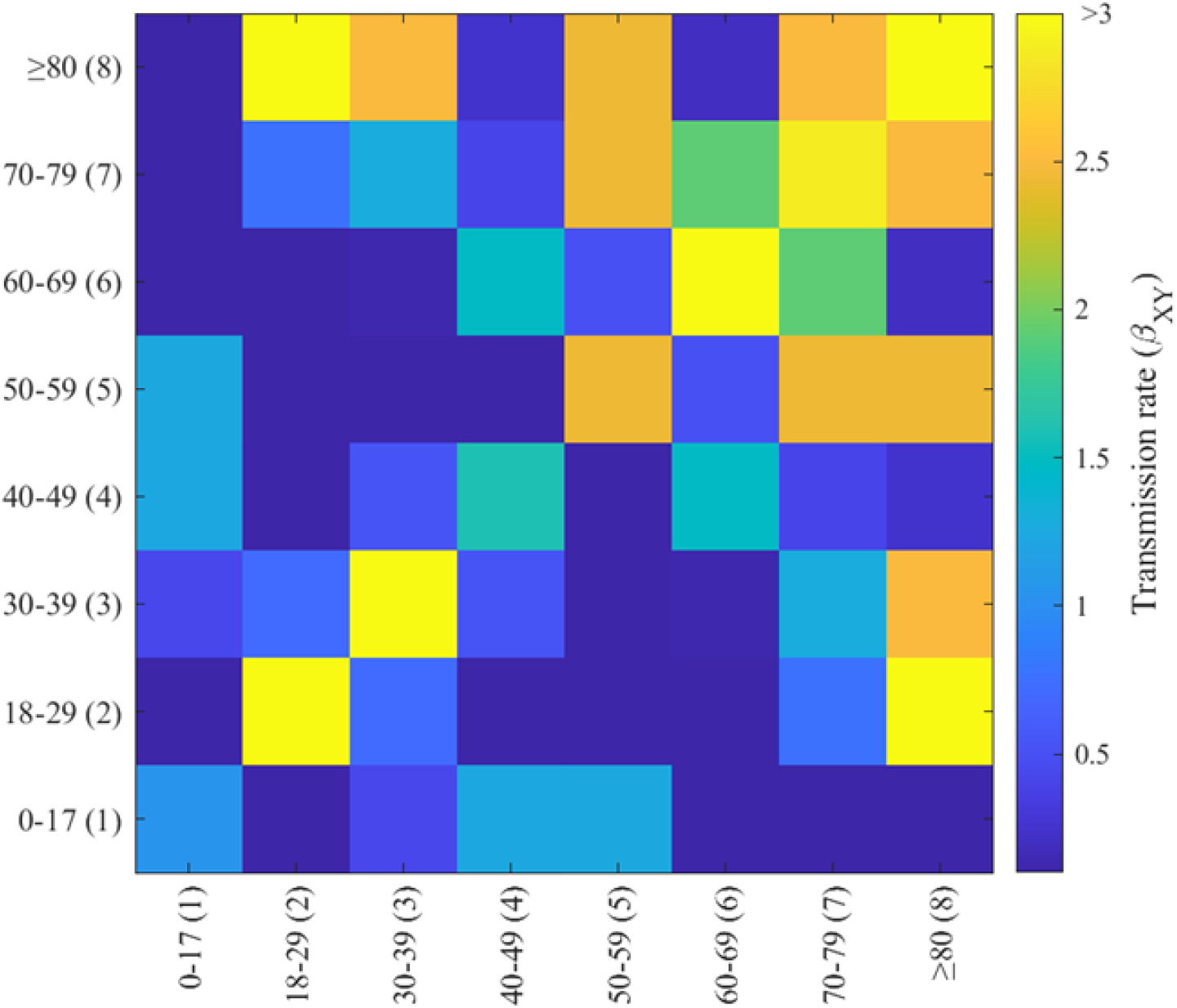
The transmission rate matrix among age groups using MLE.

### Qualification of NPIs in Korea

Fig.3 shows that the daily and cumulative incidences from the model simulation fit the data well (panel A and B, respectively). Also, the number of administered severe patients captures the trend well (panel C), even if the parameters related to severe patients were not fitted but aggregated from references.

**Fig.3.**
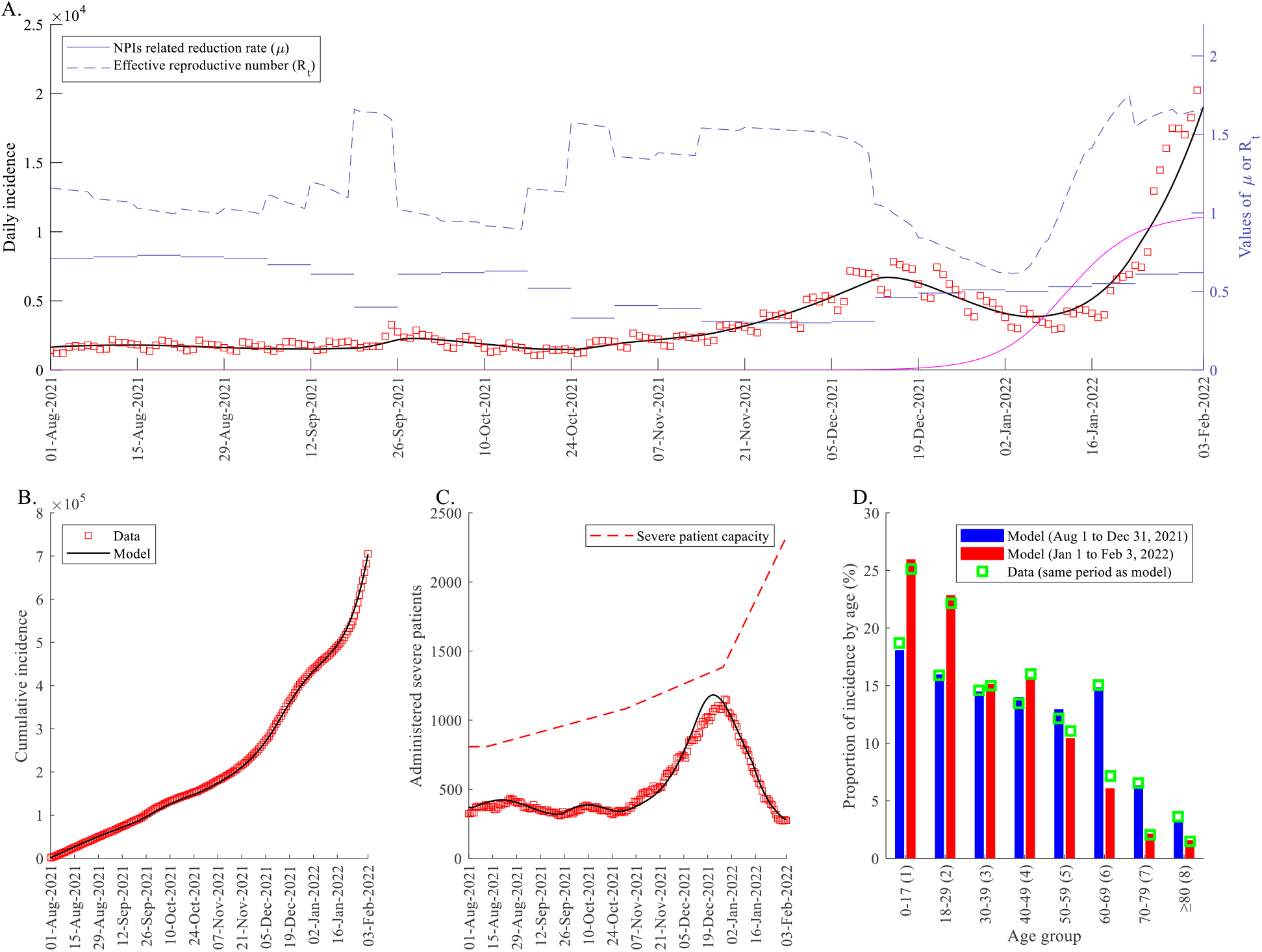
Estimation results of the qualification of nonpharmaceutical interventions (NPIs). (A) Daily incidence, NPIs-related reduction factor, and reproductive number. Dark solid curve is the model simulation and red boxes are data. Pale blue solid lines indicate NPIs-related reduction factor and dashed curve is the effective reproductive number. The magenta curve is the proportion of Omicron variant among new cases. (B) Cumulative incidence. (C) Administered severe patients. Red dashed curve indicates the severe patient capacity of Korea. (D) Proportion of incidence by age on two different periods. The blue bar is from August 1 to December 31, 2021, and the red bar is from January 1 to February 3, 2022. The green boxes indicate data.

During social distancing level 4 (August to October 2021), the range of estimated *μ* was from 0.61 to 0.73, except near the national holiday season (Chuseok, September 20 to September 22, 2021) when *μ* dropped to 0.4. Since November, as the gradual recovery policy began, *μ* decreased and ranged from 0.31 to 0.41, and later becomes 0.52 as suspended gradual recovery was announced because the number of severe patients reached more than 1,100. The obtained *μ* estimates and the corresponding values of the effective reproductive number *R*_*t*_ are illustrated as horizontal lines and dashed curves, respectively, in Fig.3 panel A. The proportion of Omicron among new cases (magenta curve) increased from 7% to 71% from December 16, 2021 to January 16, 2022 and reached 97% by the end of the simulation period.

Age groups 1 and 2 (age under 30) showed the maximum and second maximum incidence among age groups in both phases, August 1 to December 31, 2021 and January 1 to February 3, 2022, respectively. Age group 6 (60 to 69) had the third highest incidence number in 2021 but third lowest in 2022. Age groups 8 and 7 (age over 70) had the minimum and second minimum incidence during the simulation period.

### Forecast results of Omicron epidemic in 2022

Forecast from Feb 3 to the end of 2022 considering various NPIs-related reduction factor (*μ*) and maximum number of daily booster shot administration, showing the range of confirmed cases and administered severe patients, are displayed in Fig.4 If the maximum number of booster per day is 300,000 (100,000), the peak size of incidence and administered severe patients will range from 320,300 (420,900) to 1,409,200 (1,518,900) and 1,210 (1,530) to 5,120 (5,410) according to the value of *μ* which varies from 0.4 to 0.65, respectively. A secondary wave towards the end of 2022 is observed in each scenario, and the size of the secondary peak (incidence: less than 300,000, severe patient: less than 1000) is smaller than the first peak. We display the data (red boxes) until March 13, 2022, before testing policy was changed to include positive rapid antigen test done in an accredited facility as a confirmed case. ^34^

**Fig.4.**
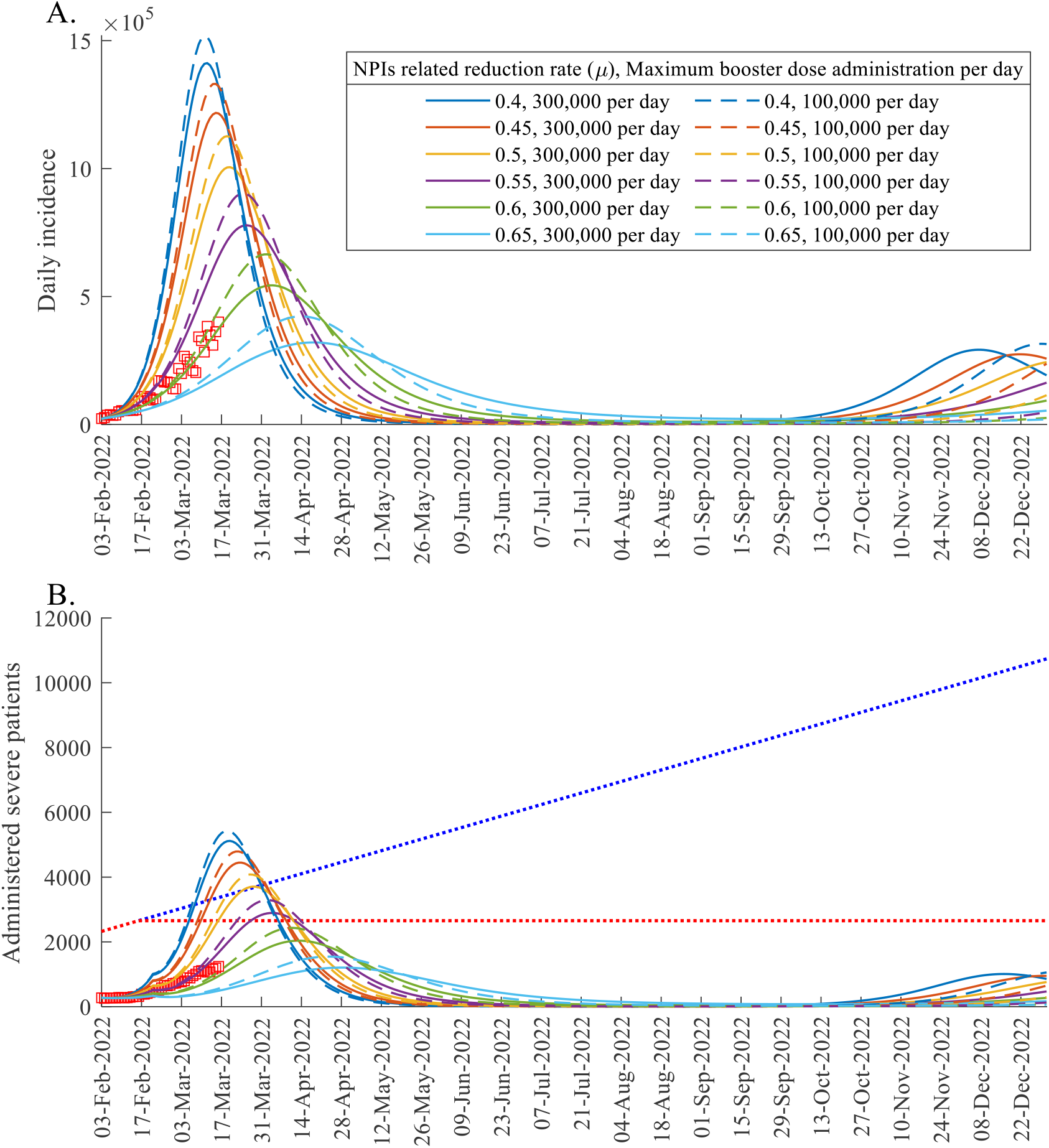
Forecast results considering different intensity of NPIs and vaccine hesitancy. (A) Daily incidence. (B) Administered severe patient. Colors of model simulation curves indicate the value of NPIs related factor (*μ*). The solid and dashed curves correspond to maximum daily booster shot administration set to 300,000 and 100,000, respectively. Blue dotted line in (B) is the expected severe patient capacity assuming that the increasing trend continues (25.42 per day, based on historical data), while the red dotted line indicates a constant trend. Red boxes are the data points.

### Time dependent impact of screening measure

Fig.5 shows the log-scaled simulation results using filled curves with different colors. Red, green, blue, and cyan areas indicate the ranges of daily incidence for the various numbers of overseas entrant cases (0.1Γ_*i*_ to 10Γ_*i*_), initiated on Nov-24, Dec-1, Dec-8, and Dec-22, respectively. As the date is delayed, the ranges of incidence become narrow. The black curve indicates the incidence when Γ_*i*_ is set to its baseline value and initiated on Nov-24. The ratio of the maximum (minimum) to the baseline incidence value when Γ_*i*_ is initiated on Nov-24, Dec-1, Dec-8, and Dec-22 are 8.64 (0.13), 2.61 (0.84), 1.36 (0.96), and 1.04 (0.99), respectively. In particular, if the impact of screening (Γ_*i*_) is varied since November 24, 2021, the range of values of the number of daily cases by February 3, 2022, is [2730, 190420]. On the contrary, if the screening is varied since December 22, 2021, the range of daily cases is [21800, 22780].

**Fig.5.**
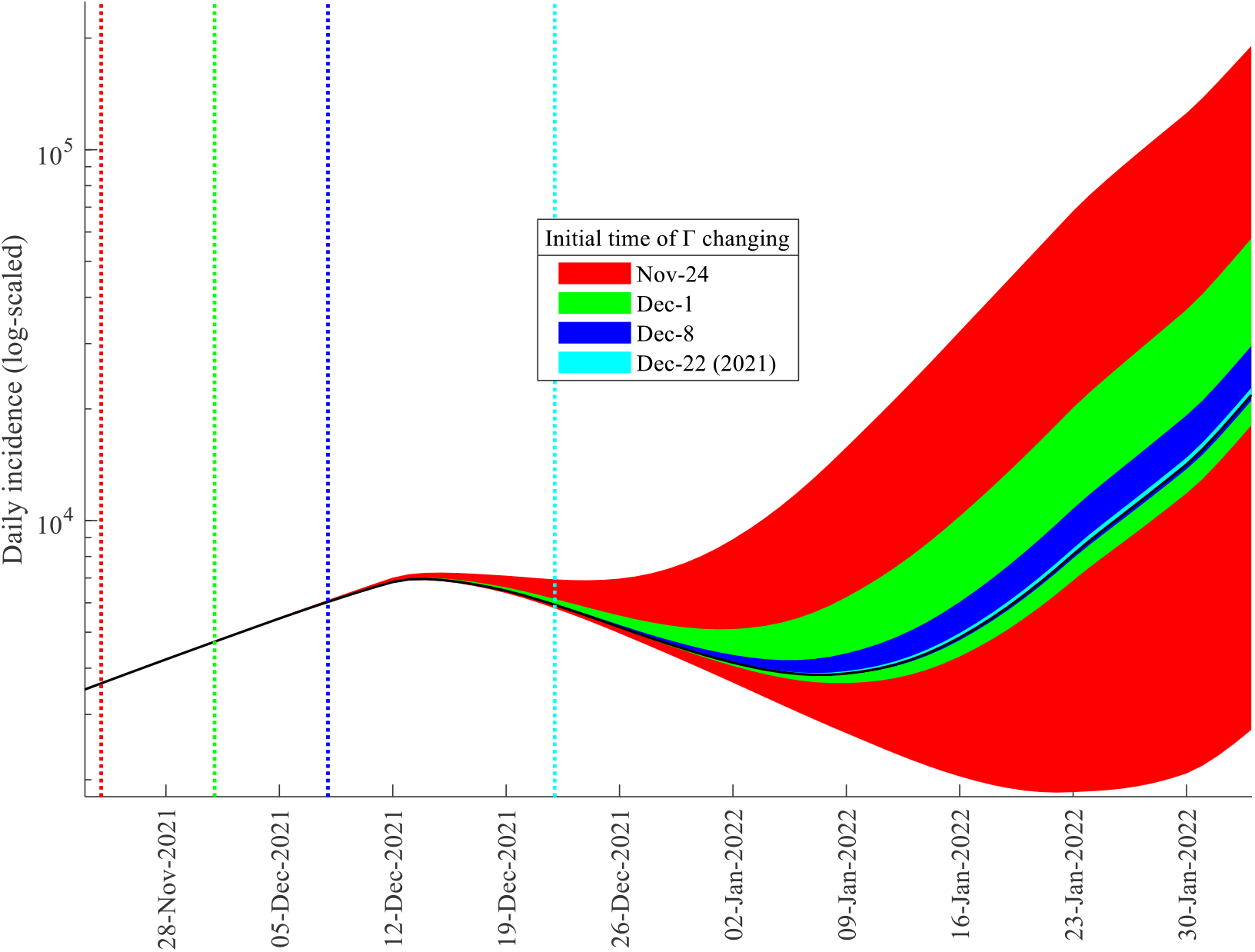
Examination of screening measure. Colored area indicates the range of daily incidence in log-scaled simulation as the number and date of daily overseas entrant case is varied.

### Endemic equilibrium study

Using Equation (1), if the severe rate, length of hospital stay, and severe patient capacity are 5%, 28 days, and 500, respectively, which might be similar to the early stage of COVID-19 in Korea, then the manageable number of incidence is approximately 360. Fig.6 illustrates the manageable daily incidence if the severe patient capacity is fixed to 2800 (panel A) or when the average length of hospital stay is set to 7 days (panel B). Panel C shows the actual incidence data and theory-driven manageable incidence using aggregated data of each day, interpolated data of hospital stay, severe rate, and severe patient capacity. ^35,36^ Blue square indicates that the manageable daily incidence of Korea in mid-February 2022 is around 600,000 assuming that severe patient capacity is 2,800, average severe rate is 0.16%, and average length of hospital stay is 7 days. Moreover, it is visible that actual daily incidence exceeded the theory-driven manageable incidence in December 2021, when the government declared the suspended gradual recovery.

**Fig.6.**
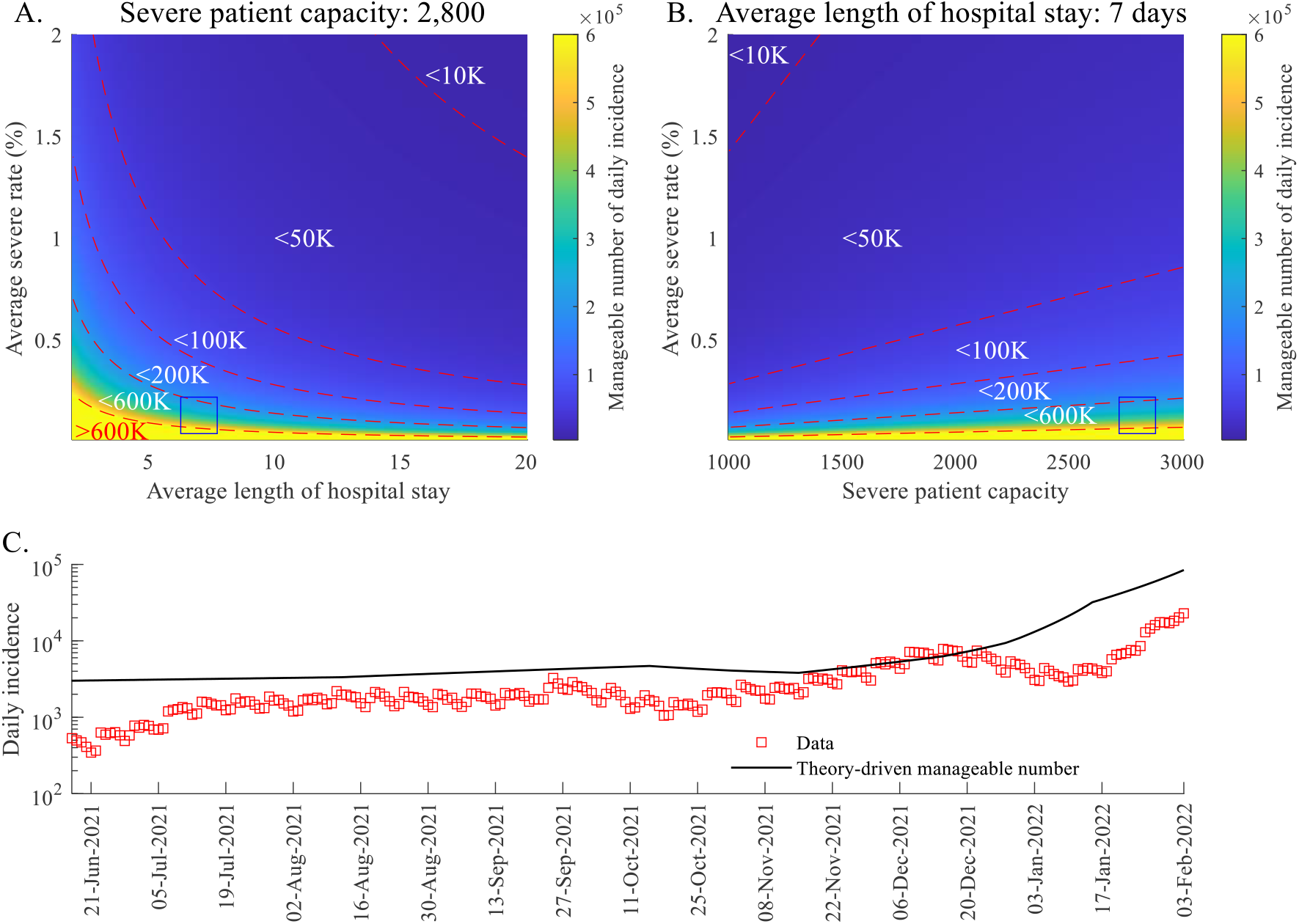
Theory-driven manageable number of daily incidence considering endemic equilibrium. (A) Color-scaled result considering varying length of hospital stay and severe rate, with fixed severe patient capacity to 2,800. Blue square indicates the severe rate of Korea in mid-February 2022. (B) Color-scaled result considering varying severe patient capacity and severe rate, with fixed length of hospital stay as 7 days. (C) Real daily incidence data and theory-driven manageable number of daily incidence using real data.

## DISCUSSION

Age-structured models are useful to analyze the heterogeneity of transmission patterns according to different age groups and suggest age-specific policies, such as vaccine prioritization or protocols related to school closures. To solve an age-structured model, transmission rate matrix (or contact matrix) is required. However, obtaining a contact matrix through survey during epidemic would be challenging. In this work, we construct the transmission matrix using MLE. The maximum value of the estimated transmission rate for age over 60 and under 60 were 6.22 and 4.06, respectively. Considering that approximately 300,000 of seniors are using elderly facilities, the transmission matrix shows the importance of disease control in elderly facilities during an epidemic. ^37^

We quantified the impact of NPIs by using *μ*, whose value was varied to indicate the different levels of social distancing policies. The range of the value of *μ* is a useful guide for the healthcare authorities in deciding the intensity of the intervention. We could also observe the risk of spreading during the holiday season, with an estimated lower *μ* value, which is a considerable factor for the policymakers. A strict social distancing, associated with high *μ* value, remains a good control measure to minimize the size of epidemic. However, there is serious economic burden if a strict policy is continued. Therefore, our model can be used as a guide in determining a more relaxed policy considering changes in the number of severe bed capacity.

In Fig.4, we observe the impact of booster shots on the number of administered severe patients under different values of NPIs-related reduction factor *μ*. In particular, it is possible for the initial peak of the green curve (*μ* =0.6) to reach the assumed value of severe patient capacity. If the maximum number of booster shots per day is small (dashed), indirectly expressing vaccine hesitancy, then the green curve reached the red-dotted line, which is a pessimistic assumption that the number of severe beds has not increased. On the other hand, the number of administered severe patients is manageable if the number of booster shots is large (solid). This result highlights the importance of booster shots in reducing the number of mild and severe infections. Finally, we observe that incidence data (red boxes) follow the green dashed curve while the administered severe patients follow the green solid curve. In the official national data, critically ill patients are defined as individuals who have SpO2 <94% on room air at sea level, a ratio of arterial partial pressure of oxygen to fraction of inspired oxygen (PaO2/FiO2) <300 mm Hg, a respiratory rate >30 breaths/min, or lung infiltrates >50%. Therefore, a patient who is infected with SARS-CoV-2 and needs ICU care for a disease other than a respiratory system problem is not defined as a critically ill patient. For this reason, data on bed use may be underestimated.

Screening measures are the primary NPIs in blocking the arrival of a new strain in the local community. However, we found that the impact of screening measures is reduced as the incoming strain becomes more dominant in the local community. Since strict screening policies incur serious socio-economic costs, strengthening screening measures might have less effect on the current situation (March 2022). Nevertheless, strengthening screening measures would be important if there is an emerging VOC outside of the country because our results suggest that strong screening measures can delay the new peak if they are applied early.

The derived formula (Eq. 1) calculates the manageable number of daily incidence cases using the data on severe rate, the average duration of hospital stay, and severe patient capacity. Factors such as emergence of relatively mild variants, vaccines, and enhanced medical support have decreased the severe rate of COVID-19 infections. Furthermore, the average duration of hospital stay reduced significantly, from 28 days to 7 days, since February 2020 to March 2022.^36,37^ The endemic equilibrium study can be useful in crafting policies that ensure the number of incidence and severe cases are within safe levels. For example, our theory-driven model indicates that the declaration of suspended gradual recovery on mid-December 2021 might have been inevitable to control the surge in daily incidence. Data on the severe patient capacity also showed a steep rise during this period (dashed curve in Fig 3 panel C).

Our mathematical-modeling-based approach is not only valid on the Delta or Omicron variants of COVID-19 but can be generally adopted for other emerging infectious disease in the future, or new variant of COVID-19. Because NPIs are incorporated using the parameter *μ*, our model would be useful as a guide in policymaking. Analysis considering various important factors, such as waning effects of vaccine and natural recovery, or variants, may give insights for the disease control. A limitation of the study is that breakthrough infection during MLE process was not considered due to the lack of available data. On March 14, 2022, confirmation of cases was expanded to include positive rapid antigen tests, which has a lower accuracy compared to PCR test. This may lead to under-reporting, which is not considered in this study. These limitations can be studied in future works.

## Data Availability

All data produced in the present study are available upon reasonable request to the authors

## ACKNOWLEDGMENTS

This paper is supported by the Korea National Research Foundation (NRF) grant funded by the Korean government (MEST) (NRF-2021M3E5E308120711). This paper is also supported by the Korea National Research Foundation (NRF) grant funded by the Korean government (MEST) (NRF-2021R1A2C100448711).

## Author Contributions

Conceptualization: Ko YS. Data curation: Ko YS, Lee JG, and Kwon DH. Formal analysis: Ko YS, Mendoza R, Mendoza V, Seo YB, Lee J, Lee JG, Kwon DH, and Jung E. Funding acquisition: Jung E. Investigation: Ko YS, Mendoza R, Mendoza V, and Jung E. Methodology: Ko YS. Software: Ko YS. Validation: Ko YS, Mendoza R, Mendoza V, and Seo YB. Visualization: Ko YS. Writing -original draft: Ko YS, Mendoza R, Mendoza V, and Jung E. Writing - review & editing: Ko YS, Mendoza R, Mendoza V, Seo YB, Lee J, Lee JG, Kwon DH, and Jung E.

