## Appendix for "Multi-faceted analysis of COVID-19 epidemic in the Republic of Korea considering Omicron variant: Mathematical modeling-based study"

### Maximum Likelihood Estimation

In this section, we introduce the parameters, transmission rates among age groups, and estimation process using the maximum likelihood estimation (MLE). To incorporate the history of infection and vaccination of individuals, we categorized the population into the following groups:

- $\Lambda_I$: infected hosts,
- $\Lambda_S$: uninfected and unvaccinated individuals,
- $\Lambda_{V-pre}$: uninfected and vaccinated before August 1, 2021, and
- $\Lambda_{V-post}$: uninfected and vaccinated after August 1, 2021.

In our likelihood estimation, we did not consider the breakthrough infection group because the available data did not contain the information whether a confirmed individual is vaccinated or not. Since we considered eight age groups (0 to 17, 18 to 29, 30 to 39, 40 to 49, 50 to 59, 60 to 69, 70 to 79, and over 80 years old), there are 32 subgroups for MLE. Individual case data, which was provided by the Korea Disease Control and Prevention Agency, contains the date of symptom onset and confirmation. We assumed that the individuals are infected 4 days before the date of symptom onset.[1] If the data on the date of symptom onset was not available, we assumed that the individual was infected 8 days (2 days of latent period + 6 days of transmission period) before.[2,3] Between two discrete time (a day), from time $t-1$ to $t$, there are two possibilities for a host in age group $i$: probability of being infected ($P_{I,i}(t)$), or not infected ($P_{S,i}(t)$). Assuming that there is a homogeneous mixing of infectors and infectees in the community and that the distribution of the infection event is exponential, then the probabilities are given by

$$P_{I,i}(t) = 1 - e^{-\sum_j \beta_{ij} \frac{I(t)}{N}},$$

$$P_{S,i}(t) = e^{-\sum_j \beta_{ij} \frac{I(t)}{N}}.$$

Note that we estimated the number of infectors ($I(t)$) by assuming that individuals can transmit the disease 2 days after they are infected until they have confirmation. To consider vaccination, we set that a vaccinated individual has partial vaccine effectiveness against infection ($0.8 \times e_i$) for rounded value of $1/\omega_i$ days, which is the average time to have vaccine-induced immunity considering secondary dose, and after $1/\omega_i$ days, a vaccinated individual has full effectiveness against infection ($e_i$). Vaccine effectiveness against infection reduces the probability of being infected ($(1 - 0.8 \times e_i) \times P_{I,i}(t)$ or $(1 - e_i) \times P_{I,i}(t)$). The parameters $\omega_i$ and $e_i$ will be described in the next subsection. Likelihood consists of the probabilities of not being infected and infected. If a host is in $\Lambda_I$, the host has $P_{S,i}$ 2 days before the host was infected and has $P_{I,i}$ a day before the host was infected and became infected. A host in the other groups only has $P_{S,i}$ until the final time of MLE. A more detailed explanation of MLE process can be found in a previous work.[4]

The initial time of MLE is August 1, 2021, and the final time is set to December 31, 2021. During this period, the Delta variant was dominant. Therefore, the estimated matrix $M_1$ represents the transmission dynamics of the Delta variant. However, $M_1$ contains the effect of NPIs. Hence, the matrix is adjusted to exclude this effect so that we can incorporate NPIs as a separate parameter $\mu$ in the mathematical model. The adjusted matrix $M_2$ is calculated as $M_2 = M_1 \frac{R_0^\delta}{R_{est}}$, where $R_{est}$ is the reproductive number calculated using $M_1$ and $R_0^\delta = 6.22$ is the basic reproductive number of the Delta variant.[5,6]

**Mathematical modeling of COVID-19 considering Delta and Omicron variants**

The mathematical formulation of the model is as follows:

$$X \in \{u, w, v1, v2, wv, b, wb\}, \qquad i = 1, \ldots, 8,$$

$$\frac{dX_i}{dt} = -\lambda_{X,i}^{\delta} X_i - \lambda_{X,i}^{o} X_i - OUT_{X,i} + IN_{X,i},$$

$$\frac{dE_{X,i}^{\delta}}{dt} = \lambda_{X,i}^{\delta} X_i - \kappa E_{X,i}^{\delta}, \qquad \frac{dE_{X,i}^{o}}{dt} = \Gamma_i + \lambda_{X,i}^{o} X_i - \kappa E_{X,i}^{o},$$

$$\frac{dI_{X,i}^{\delta}}{dt} = \kappa E_{X,i}^{\delta} - \alpha I_{X,i}^{\delta}, \qquad \frac{dI_{X,i}^{o}}{dt} = \kappa E_{X,i}^{o} - \alpha I_{X,i}^{o},$$

$$\frac{dQ_{X,i}^{\delta}}{dt} = \alpha I_{X,i}^{\delta} - \sigma Q_{X,i}^{\delta}, \qquad \frac{dQ_{X,i}^{o}}{dt} = \alpha I_{X,i}^{o} - \sigma Q_{X,i}^{o},$$

$$\frac{dM_{X,i}^{\delta}}{dt} = \left(1 - p_{X,i}^{\delta}\right)\sigma Q_{X,i}^{\delta} - \gamma_m^{\delta} M_{X,i}^{\delta}, \qquad \frac{dM_{X,i}^{o}}{dt} = (1 - p_{X,i}^{o})\sigma Q_{X,i}^{o} - \gamma_m^{o} M_{X,i}^{o},$$

$$\frac{dC_{X,i}^{\delta}}{dt} = p_{X,i}^{\delta}\sigma Q_{X,i}^{\delta} - \gamma_c^{\delta} C_{X,i}^{\delta}, \qquad \frac{dC_{X,i}^{o}}{dt} = p_{X,i}^{o}\sigma Q_{X,i}^{o} - \gamma_c^{o} C_{X,i}^{o},$$

$$\frac{dR_{X,i}}{dt} = \gamma_m^{\delta} M_{X,i}^{\delta} + \gamma_m^{o} M_{X,i}^{o} + \left(1 - f_{X,i}^{\delta}\right)\gamma_c^{\delta} C_{X,i}^{\delta} + \left(1 - f_{X,i}^{o}\right)\gamma_c^{o} C_{X,i}^{o} - \zeta_X R_{X,i},$$

$$\lambda_{X,i}^{\delta} = (1 - \mu)\left(1 - e_{X,i}^{\delta}\right)\sum_X \sum_j \beta_{ij}\left(1 - \widetilde{e_{X,j}^{\delta}}\right) I_{X,j}^{\delta},$$

$$\lambda_{X,i}^{o} = (1 - \mu)\left(1 - e_{X,i}^{o}\right)\sum_X \sum_j \beta_{ij}(1 - \widetilde{e_{X,j}^{o}}) I_{X,j}^{\delta},$$

$$OUT_{u,i} = \min\left(u_i, v_i(t)\right), \qquad IN_{u,i} = 0,$$

$$OUT_{w,i} = v_i(t) - \min\left(u_i, v_i(t)\right), \qquad IN_{w,i} = \zeta(R_{u,i} + R_{w,i}),$$

$$OUT_{v1,i} = \omega_i v1_i, \qquad IN_{v1,i} = v_i(t),$$

$$OUT_{v2,i} = \tau_{v,i} v2_i + b_i(t) - \min\left(wv_i, v_i^b(t)\right), \qquad IN_{v2,i} = \omega_i v1_i,$$

$$OUT_{wv,i} = \min\left(wv_i, v_i^b(t)\right), \qquad IN_{wv,i} = \tau_v v2_i + \zeta_v(R_{v1,i} + R_{v2,i} + R_{wv,i}),$$

$$OUT_{b,i} = \tau_b b_i, \qquad IN_{b,i} = b_i(t),$$

$$OUT_{wb,i} = 0, \qquad IN_{wb,i} = \tau_b b_i + \zeta_v(R_{b,i} + R_{wb,i}),$$

where $\mu$ indicates the reduction rate caused by NPIs. For example, ignoring other factors, if the basic reproductive number is 2 and $\mu$ is 0.7, then the effective reproductive number becomes $(1-0.7)\times 2=0.6$. We estimated the value of $\mu$ every week using least square curve fitting method, minimizing the difference of cumulative incidence comes from model ($\int \sum_X \sum_i \alpha(I^{\delta}_{X,i}+I^{o}_{X,i}))\,dt$) and data. Model simulation time was from 1 August 2021 to 2 February 2022, because the testing policy has been changed since 3 February 2022.[7] The parameter $\Gamma_i$ represents the number of overseas entrants cases from age group $i$ who are not screened but entered the local community. Then using the population data of the age groups, $\Gamma_i$ is computed using ratio and proportion and using the formula $40/38=\sum_X \sum_i \Gamma_i$, where 40/38 is the average daily number of overseas entrants cases (40 in total) across all ages from 24 November to 31 December 2021 (38 days). The model parameters are listed in Table[a] and Table[b]. Note that there are three effectiveness of vaccine, against infection, transmission, and severity. Daily number of vaccine administration, $v_i^b(t)$ and $v_i(t)$, are obtained from data.[8] In Korea, booster shots were administered to anyone who finished primary doses more than 90 days ago. That means, targets for the booster shot administration are waned after primary dose ($wv_i$) and 2 weeks after primary dosed ($v2_i$). Therefore, to keep nonnegativity condition and prioritize waned hosts to be boostered, number of flows from $wv_i$ and $v2_i$ to being boostered ($b_i$) are set as $\min\left(wv_i, v_i^b(t)\right)$ and $v_i^b(t)-\min\left(wv_i, v_i^b(t)\right)$, respectively.

Table[a] Model parameters, non-age dependent

| Symbol | Description | Value | Reference |
|---|---|---|---|
| $1/\kappa$ | Latent period | 2 | [1,3] |
| $1/\alpha$ | Infectious period | 6 | [2,3] |
| $\sigma$ | Progression rate | 2 | Assumed |
| $\gamma_m^\delta$ | Recovery rate of mild Delta case | 1/10 | [9] |
| $\gamma_c^\delta$ | Recovery rate of severe Delta case | 1/14 | [9] |
| $\gamma_m^o$ | Recovery rate of mild Omicron case | 1/7 | [9] |
| $\gamma_c^o$ | Recovery rate of severe Omicron case | 1/7 | [9] |
| $\zeta_u, \zeta_w$ | Waning rate of infection-induced immunity for unvaccinated case | 1/120 | [10,11,12] |
| $\zeta_{v1}, \zeta_{v2}, \zeta_{wv}, \zeta_b, \zeta_{wb}$ | Waning rate of infection-induced immunity for vaccinated case | 1/480 | [13] |
| $\tau_b$ | Waning rate of booster shot | 1/300 | Assumed |

Table[b] Model parameters, age-and-strain dependent. Note that subscript and superscript indicate strain and age group, respectively

| Symbol | Description | 1 | 2 | 3 | 4 | 5 | 6 | 7 | 8 | Reference |
|---|---|---|---|---|---|---|---|---|---|---|
| $1/\omega_i$ | Average duration from first dose to 2 weeks after second dose | 35 | 38 | 39 | 43 | 43 | 86 | 66 | 41 | 14 |
| $\tau_{v,i}$ | Waning rate after finishing primary vaccine | 0.0020 | 0.0021 | 0.0025 | 0.0023 | 0.0023 | 0.0036 | 0.0030 | 0.0022 | 8,15,16 |
| $p_{u,i}^{\delta}$ | Severe rate of unvaccinated | 0.0004 | 0.0030 | 0.0087 | 0.0200 | 0.4323 | 0.0749 | 0.1329 | 0.2587 | 17 |
| $p_{u,i}^{o}$ | | 0.0001 | 0.0008 | 0.0022 | 0.0050 | 0.0106 | 0.0187 | 0.0332 | 0.0647 | |
| $p_{v1,i}^{\delta}, p_{v2,i}^{\delta}, p_{wv,i}^{\delta}, p_{b,i}^{\delta}, p_{wb,i}^{\delta}, p_{w,i}^{\delta}$ | Severe rate, except unvaccinated | 0 | 0 | 0.0038 | 0.0051 | 0.0080 | 0.0339 | 0.0244 | 0.0541 | 17 |
| $p_{v1,i}^{o}, p_{v2,i}^{o}, p_{wv,i}^{o}, p_{b,i}^{o}, p_{wb,i}^{o}, p_{w,i}^{o}$ | | 0 | 0 | 0.0009 | 0.0013 | 0.0020 | 0.0085 | 0.0061 | 0.0135 | |
| $f_{X,i}^{\delta}, f_{X,i}^{o}$ | Fatal rate | 0 | 0.0001 | 0.0003 | 0.0006 | 0.0020 | 0.0064 | 0.0254 | 0.1275 | 17 |
| $e_{w,i}^{\delta}$ | Natural-infection-induced partial immunity against infection | 0.3960 | 0.4147 | 0.3818 | 0.3938 | 0.4015 | 0.2737 | 0.3226 | 0.3810 | 8,15,16 |
| $e_{w,i}^{o}$ | | 0.0710 | 0.0765 | 0.0596 | 0.0656 | 0.0691 | 0.0064 | 0.0323 | 0.0630 | 8,15,16 |
| $e_{v1,i}^{\delta}$ | Vaccine effectiveness against infection; 2 weeks before finishing primary doses | 0.6216 | 0.6313 | 0.6109 | 0.6183 | 0.6228 | 0.5450 | 0.5757 | 0.6122 | 8,15,16 |
| $e_{v1,i}^{o}$ | | 0.2320 | 0.2437 | 0.1888 | 0.2079 | 0.2187 | 0.0204 | 0.1053 | 0.2058 | 8,15,16 |
| $e_{v2,i}^{\delta}$ | Vaccine effectiveness against infection; 2 weeks after finishing primary doses | 0.7770 | 0.7891 | 0.7636 | 0.7728 | 0.7785 | 0.6812 | 0.7196 | 0.7652 | 8,15,16 |
| $e_{v2,i}^{o}$ | | 0.2900 | 0.3046 | 0.2360 | 0.2599 | 0.2734 | 0.0255 | 0.1317 | 0.2572 | 8,15,16 |
| $e_{wv,i}^{\delta}, e_{wb,i}^{\delta}$ | Waned vaccine effectiveness against infection | 0.3960 | 0.4147 | 0.3818 | 0.3938 | 0.4015 | 0.2737 | 0.3226 | 0.3810 | 8,15,16 |
| $e_{wv,i}^{o}, e_{wb,i}^{o}$ | | 0.0710 | 0.0765 | 0.0596 | 0.0656 | 0.0691 | 0.0064 | 0.0323 | 0.0630 | 8,15,16 |
| $e_{b,i}^{\delta}$ | Vaccine effectiveness against infection; boostered | 0.8990 | 0.8962 | 0.8991 | 0.8980 | 0.8972 | 0.9094 | 0.9053 | 0.9003 | 8,15,16 |
| $e_{b,i}^{o}$ | | 0.5100 | 0.5012 | 0.4962 | 0.4975 | 0.4975 | 0.4869 | 0.4963 | 0.5070 | 8,15,16 |
| $\widetilde{e_{w,i}^{\delta}}$ | Natural-infection-induced transmissibility reduction | 0.0500 | 0.0547 | 0.0503 | 0.0520 | 0.0532 | 0.0348 | 0.0408 | 0.0482 | 8,15,16 |
| $\widetilde{e_{w,i}^{o}}$ | | 0.0060 | 0.0065 | 0.0051 | 0.0056 | 0.0059 | 0.0005 | 0.0027 | 0.0053 | 8,15,16 |
| $\widetilde{e_{v1,i}^{\delta}}$ | Vaccine-induced transmissibility reduction; 2 weeks before finishing primary doses | 0.2160 | 0.2338 | 0.2256 | 0.2292 | 0.2325 | 0.1908 | 0.2007 | 0.2130 | 8,15,16 |
| $\widetilde{e_{v1,i}^{o}}$ | | 0.0320 | 0.0350 | 0.0273 | 0.0300 | 0.0317 | 0.0029 | 0.0146 | 0.0284 | 8,15,16 |
| $\widetilde{e_{v2,i}^{\delta}}$ | Vaccine-induced transmissibility reduction; 2 weeks after finishing primary doses | 0.2160 | 0.2338 | 0.2256 | 0.2292 | 0.2325 | 0.1908 | 0.2007 | 0.2130 | 8,15,16 |
| $\widetilde{e_{v2,i}^{o}}$ | | 0.0320 | 0.0350 | 0.0273 | 0.0300 | 0.0317 | 0.0029 | 0.0146 | 0.0284 | 8,15,16 |
| $\widetilde{e_{wb,i}^{\delta}}, \widetilde{e_{wv,i}^{\delta}}$ | Waned vaccine- | 0.0500 | 0.0547 | 0.0503 | 0.0520 | 0.0532 | 0.0348 | 0.0408 | 0.0482 | 8,15,16 |

| | | | | | | | | | | |
|---|---|---|---|---|---|---|---|---|---|---|
| $\widetilde{e^{o}_{wb,\iota}}, \widetilde{e^{o}_{wv,\iota}}$ | induced transmissibility reduction | 0.0060 | 0.0065 | 0.0051 | 0.0056 | 0.0059 | 0.0005 | 0.0027 | 0.0053 | [8,15,16] |
| $\widetilde{e^{\delta}_{b,\iota}}$ | Vaccine-induced transmissibility reduction; boostered | 0.4100 | 0.4026 | 0.4044 | 0.4035 | 0.4024 | 0.4142 | 0.4126 | 0.4104 | [8,15,16] |
| $\widetilde{e^{o}_{b,\iota}}$ | | 0.0770 | 0.0747 | 0.0740 | 0.0741 | 0.0740 | 0.0734 | 0.0749 | 0.0765 | [8,15,16] |
